# Approach in Inputs & Outputs Selection of Data Envelopment Analysis (DEA) Efficiency Measurement in Hospital: A Systematic Review

**DOI:** 10.1101/2023.10.18.23297223

**Authors:** M Zulfakhar Zubir, A Azimatun Noor, AM Mohd Rizal, A Aziz Harith, M Ihsannuddin Abas, Zuriyati Zakaria, Anwar Fazal A.Bakar

## Abstract

Data Envelopment Analysis (DEA) has been employed as a performance evaluation tool in the evaluation of efficiency and productivity in numerous fields. This includes hospitals in particular as well as the broad healthcare industry. This review examines 89 papers that discuss the use of DEA in hospitals, paying particular attention to approaches for choosing inputs and outputs as well as the most recent developments in DEA studies. English articles with empirical data from year 2014-2022 (Web of Science, Scopus, PubMed, ScienceDirect, Springer Link, and Google Scholar) were extracted based on PRISMA methodology. DEA Model parameters were specified based on previous studies and approaches were identified narratively. The approaches can be grouped into four: (1) Literature review, (2) Data availability, (3) Systematic method and (4) Expert judgement. The approaches were applied as one strategy either by itself or in combination with others. This review’s emphasis on approaches used in hospital may constrain its conclusions. There might be another strategy or method used to select the input and output for a DEA study in a different area or strategies based on different viewpoints. The trend for DEA application were quite similar to previous studies. There is no evidence that one model fits all DEA model parameters better than another. Based on the reviewed literature, we offer some recommendations and methodological principles for DEA studies.

## Introduction

Efficiency is a term that does not need to be introduced in the world of economics. Following the seminal works of Farrell in 1957, which showed the measurement of efficiency as one that accounts for all inputs and outputs while avoiding index number issues and demonstrating how it may be calculated in practice. Indeed, the studies of efficiency gained much interest not only from statisticians and economists but a wider range audience [1]. These include those in the healthcare and medicine areas. There is no consensus on the best method to measure efficiency. Measurement techniques such as Data Envelopment Analysis (DEA), Stochastic Frontier Analysis (SFA), Pabon Lasso, and Ratio Analysis are among the various methodologies used for efficiency studies in health facilities [2–4]. In the Global Programme on Evidence for Health Policy Discussion Paper Series by World Health Organization (WHO) unveiled their own method for evaluating the effectiveness of healthcare systems. They explicitly specified the broad set of goals of the health system, including responsiveness (both level and distribution), fair finance, and health inequality, in addition to the more conventional goal of population health, in contrast to past work in this field [5]. While in another report, the WHO measures the health system’s performance by introducing a measure of national health systems’ success in attempting to meet three broad goals: good health, responsiveness to the population’s expectations, and fairness of financial contribution [6]. Even with that, there is controversy and critique on the method used while there is an agreement on making those assessments correctly focus, analyse more critically, more positively, and aid in a crucial conversation between health systems stakeholders [7–9]. In a more recent years, DEA has been used to measure the efficiency of 180 countries based on six key dimensions of healthcare systems namely clinical outcomes, health adjusted life years, access, equity, safety, and resources [10].

Thus, the stakeholders must understand there cannot be efficiency metrics that are universally applicable and appropriate for all healthcare systems. It is vital to have a thorough grasp of the institutional arrangements, data, and measurements in order to choose suitable measures, resources, and other parts of the health system, and a framework is designed. in keeping with the analysis. It should be noted that the optimal method to apply performance measurement is not to determine the one simple adjustment that can be made in a supporting role to enhance one of the health system outcomes, but to employ it as a means of gauging performance more broadly among the various system components [11,12].

Numerous indicators, ranging from measures that compare activities to measures that compare expenses, are available to determine if limited resources for health are being used most effectively. The use of quantitative metrics in evaluating hospital performance is primarily of interest. Additionally, a wide variety of indicators were utilized to assess the quality of hospital services [13,14]. Efficiency comparisons can be transparently compared using techniques based on strong economic theory. Two of the most popular techniques for calculating the effectiveness of health care are data envelopment analysis (DEA) and stochastic frontier analysis (SFA) [11]. Over the past 40 years since the earliest study by Nunamaker in 1983, these techniques have been applied extensively in published applications in healthcare settings [15–19]. Especially DEA despite the known theoretical and methodological limitations had gained interest from researchers to address these limitations and development of multiple methods which integrate DEA with other statistical techniques and methodologies to improve efficiency evaluation [20,21].

### 1.1 DEA as an efficiency analysis tool in hospital

DEA is a mathematical technique for assessing the relative efficiency of homogenous decision-making units (DMUs) with many inputs and numerous outputs. It was originally created within the operations research and econometrics disciplines. The disadvantage of DEA is that it is non-parametric and deterministic, which indicates that outliers are more noticeable. The effectiveness of a DMU is then evaluated in comparison to the effectiveness of every other member of the group: The most effective DMUs are those at the efficiency frontier, which have maximum outputs produced by using the same level of inputs as all other DMUs [17,22]. In the DEA literature, this is a well-known CCR model (Charnes, Cooper, and Rhodes) or Constant Return to Scale (CRS) assumption as the input-output correlation can be examined even in the absence of congestion effects of any kind. In other words, the output may increase in a precise linear relationship with the inputs [10,23]. This model and assumption were further extended by Banker with the BCC model (Banker, Charnes, and Cooper) and variable returns to scale (VRS) assumption which assumes that economies’ scales shift as the DMU’s size grows [23,24].

Aside from the model type and return to scale assumption, DEA also considers the model orientation either input-oriented or output-oriented. The assumption behind input orientation is that a DMU can control more inputs than outputs. The opposite argument, however, might be made: that given their ability for inputs to raise their organization’s efficiency, the organizations can increase their outputs [23,25]. When calculating a DMU’s or an organization’s efficiency using DEA, it’s critical to note the inputs and outputs. The selection and combination of both must be precise, thorough, pertinent, and appropriate in order to accurately depict the hospital’s functions and satisfy the needs of stakeholders who are evaluating the hospital’s efficiency [18,21]. In addition to this, advanced analysis had been incorporated in DEA which includes further models of CCR and BCC, longitudinal or window analysis including The Malmquist index, statistical analysis such as regression and bootstrapping method [20,23,25–27].

### 1.2 Input and output selection in hospital DEA application

Numerous studies have been published to demonstrate the usefulness and potential of DEA in evaluating hospital efficiency[28–31]. Although creating hypotheses by comparing DEA ratings across several hospital studies is helpful, there are significant drawbacks.

- varying input and output metrics throughout various time frames
- the DEA score distribution is so skewed that relying on the standard measures of central tendency will be inaccurate
- the study’s output metrics diverge greatly from one another
- hospital production models and types that differ greatly

However, there have been creative and innovative studies conducted at the hospital level that may be helpful to decision-makers [23,32]. The methodology of DEA has been the subject of dozens of review studies on hospital application. General applications of DEA to health care performance measurement[15,16,18], categorization or clustering of DEA technique [20,33], comparing DEA analysis between other methods, between countries or between two periods [28,30,34,35], or new knowledge and new approach on the assessment of DEA [17,21].

A systematic literature review (SLR) uses organized, transparent, and replicable techniques at each stage of the process to completely discover and synthesize literature that is relevant to a certain subject. It adheres to a protocol (detailed plan) that predetermines its main goals, ideas, and techniques.

Decisions and actions are meticulously recorded so that readers can follow them and assess the reviewer’s methodology [36]. Despite the large number of studies on the use of DEA in hospitals, efforts to comprehensively examine these studies are still missing and warrant more investigation. This article identifies and describes DEA research in hospitals in an effort to close the knowledge gap.

What is the approach to choose the optimal input and output for measuring DEA efficiency is the major research topic that served as the foundation for the current article’s systematic literature review? This study’s primary emphasis is on the approach or way of selecting the input and output adopted by researchers in the application of DEA. Hospitals received special attention because they are among the institutions whose efficiency is hardest to gauge because of their dynamic nature of service production and variation across providers [25,37,38]. To the best of our knowledge, no research has previously looked into or investigated the approach in selecting input and output with a focus on DEA hospital applications. Due to this, we carried out this research. In addition, this study observed the current trend in analysing hospital efficiency using DEA.

The remainder of this essay is structured as follows: The methodology section and the PRISMA Statement (Preferred Reporting Items Systematic Reviews and Meta-Analysis) methods are described in the second section. The scientific literature is methodically reviewed and synthesised in the third section, and the approach used to select the input and output for DEA in hospitals are examined and evaluated in the fourth section. Limitation and conclusion are outlined in the final section.

## Methodology

This section discusses the process used to find articles on DEA that are relevant to gauging hospital efficiency. We employed the PRISMA methodology, which consists of resources (Web of Science, Scopus, PubMed, ScienceDirect, Springer Link, and Google Scholar) used to conduct the SLR, eligibility and exclusion criteria, review process steps (identification, screening, eligibility), and data abstraction and analysis.

### 2.1 PRISMA

PRISMA is a minimal set of elements for reporting in systematic literature reviews and meta-analyses that are supported by evidence. PRISMA is primarily concerned with reporting reviews that assess the effects of interventions, but it can also serve as a foundation for reporting SLR with goals other than assessing interventions [39]. Future researchers should be provided with a comprehensive manual on the SLR methodological approach. The first step in SLR is to create and validate the review procedure, publication standard, and reporting standard/guidance, which are manuals of systematic plans that direct researchers on what should be taken into account throughout the review [40].

### 2.2 Journal databases

Six databases were used to search for articles published from 2014 until 2022 and these were Web of Science, Scopus, PubMed, ScienceDirect, Springer Link, and Google Scholar. There are noticeable performance variations across the search engines analysed, demonstrating that there is no ideal search method. Their effective usage consequently necessitates that searcher are well-trained, can assess a system’s strengths and shortcomings, and can decide where and how to search based on that information. We chose the six databases because they may offer a carefully curated medical database with features and tools that help with recall and a wide range of options to maximize precision [41,42].

### 2.3 Identification

The systematic review procedure included four steps. During the first stage, search terms were identified. We performed a search based on prior research using the terms “efficiency*”, “performance*”, “productivity*”, “benchmark*”, “hospital*”, “data envelopment analysis”, and “DEA”.

### 2.4 Screening

We established inclusion and exclusion criteria. To start, only journals with articles containing empirical data are chosen, therefore review articles (SLR and SR), book series, books, chapters in books, and conference proceedings are all disregarded. Second, the search attempts excluded non-English publications and concentrated only on items written in English, avoiding any ambiguity or difficulty in translation. Thirdly, in terms of the chronology, a period of 9 years is chosen (between 2014 and 2022), which is long enough to see changes in research and publications that are connected. The period selected also aims as a continuation of the previous study by O’Neill et al. (1984 to 2004), Cantor & Poh (1994 to 2017), and Kohl et al. (2005 to 2016). The final number of articles for the quality appraisal stage were 89. A PRISMA (Fig 1) diagram provides a detailed description of the entire search procedure.

**Fig 1.**
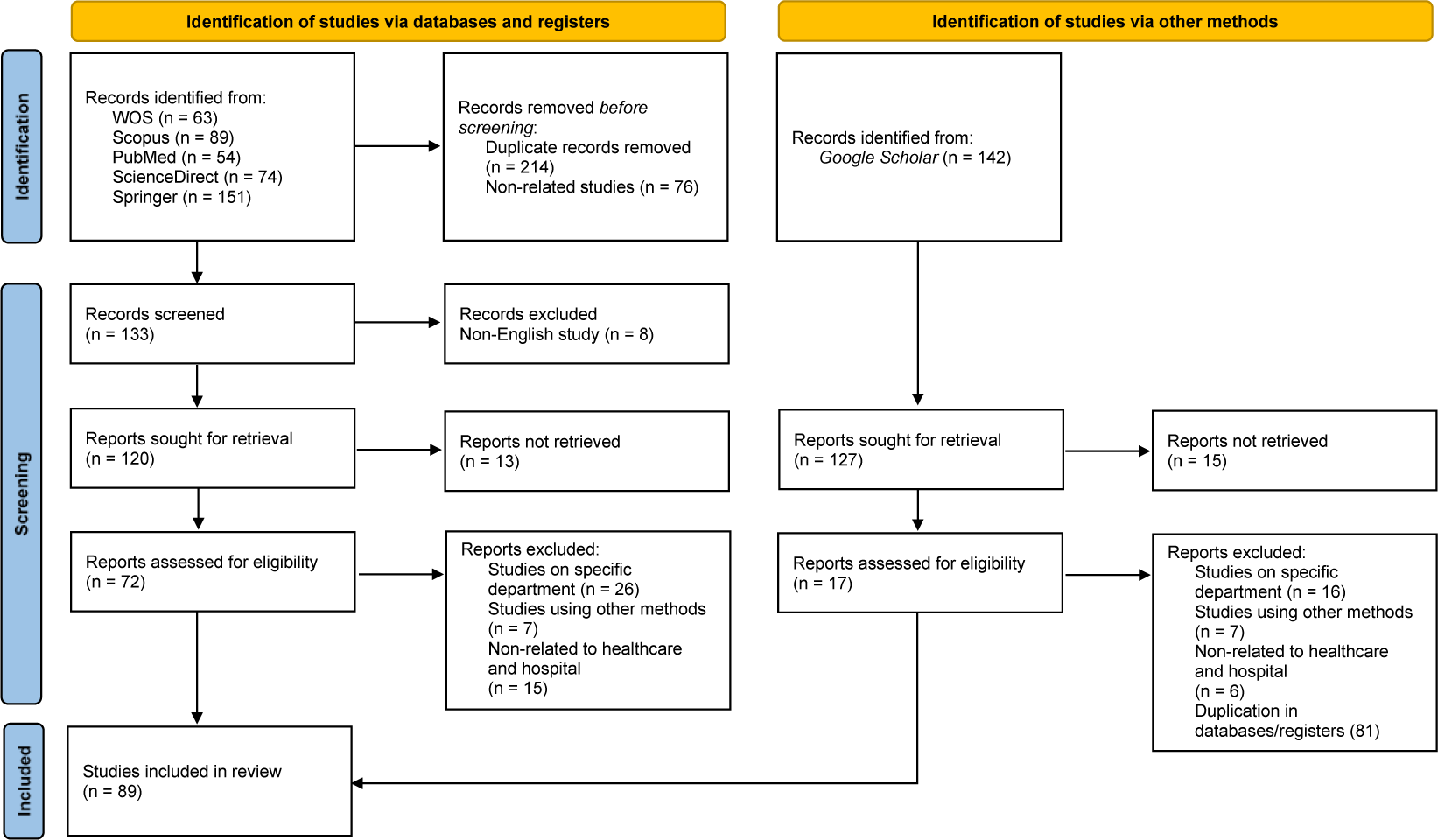
PRISMA diagram of the search process.

### 2.5 Quality appraisal

To make sure that the methodology and analysis of the chosen studies were done in a satisfactory manner, the quality appraisal stage was carried out. For this purpose, we used two quality appraisals tools that cover knowledge transfer [35,43] and economic evaluations and efficiency measurement [44,45]. The fifteen-point scale developed by Mitton et al. covered the following topics: evaluation of the literature and identifying research gaps; research question and design, validity, and reliability; data collecting; population and sampling; and analysis and reporting of results. These criteria were given a score of 0 for not being present or reported, 1 for being present but of low quality, 2 for being present and of mid-range quality, or 3 for being present and of high quality [43]. While The four dimensions covered by the Varabyova and Müller checklist were reporting, external validity, bias, and power. The quality assessment checklist’s items were all given a score of 0 (no/unclear) or 1. (yes). One item was specific to research using a second-stage analysis since it addressed the potential sources of bias in the study. Studies that did not undertake a second-stage analysis received a maximum score of 13, while studies that did receive a second-stage analysis received a maximum score of 14. Only the items relevant to that study’s design were used to determine the maximum score (100%) for each study [45].

We are not aware of any such established standards for evaluating the planning or execution of research on healthcare efficiency indicators. Thus, in order to be more robust and minimize bias, we choose to evaluate the scientific soundness of the chosen research using two tools. For more reliability, all of the chosen publications were separately evaluated by two co-authors from different institutions using both tools. In the event of a disagreement, a third reviewer was asked to evaluate the work.

### 2.6 Data extraction and analysis

The articles underwent evaluation and analysis. Focused efforts were made on particular studies that addressed the study purpose. Reading the abstracts first, then the entire articles, allowed for the data extraction. (in-depth). To determine the approach in selecting input and output for hospital DEA study, content analysis was used in conjunction with quantitative and qualitative analysis. A data extraction form with entries for year of publication, country of study, type of hospitals studied, number of hospitals, number of observations (DMUs), model types, return to scale, model orientation, type of efficiency measured, inputs and outputs, number of models tested, application of second stage analysis in the study, and approach used in selecting inputs/outputs, was used to collect data from the selected studies.

### 2.7 Statistical analysis

The agreement between two raters (co-authors) in evaluating the studies was measured using Intra-class correlation (ICC). By comparing the variability of various evaluations of the same subject to the overall variation across all ratings and all subjects, ICC evaluates the dependability of ratings. The evaluations are quantitative. The ICC coefficient value for the Mitton et al.’s fifteen-point scale and Varabyova and Müller economic evaluations and efficiency measurement are 0.956 and 0.984 respectively. No articles were eliminated at this point because the review was in qualitative and quantitative, however articles with high quality ratings were given more consideration in the data analysis and result interpretation.

## Results

Eighty-nine papers that met the criteria for inclusion were all retrospective studies that were published between 2014 and 2022. A summary of all the specifics of the listed research may be found in Appendix A and Appendix B.

### 3.1 Efficiency analysis

Efficiency analysis in DEA focus on the data by measuring the performance set of DMUs. The definition of DMU is generic and broad. In this review it focuses on the “hospital”. Technical efficiency, scale efficiency, pricing efficiency, and allocative efficiency are the four main ideas of efficiency [25]. While other describe efficiency as technical, pure, scale, allocative, cost, and congestion. DEA able to perform efficiency analysis at single point of time and over time [26]. Thus, the data can be cross sectional (single period) or longitudinal (panel data). In longitudinal analysis DEA measure efficiency by two methods which are, Malmquist Productivity Index (MPI) and Window Analysis (WA).

The majority of the papers that were included (29 of 89, 32.58%), evaluated hospital performance solely on the basis of Pure Technical Efficiency (PTE) [46–74]. Pure Technical Efficiency (PTE) is the effectiveness with which an input set is utilised to produce an output on the VRS frontier [50,69]. In other words, a hospital is considered technically efficient if it generates the greatest amount of output from the fewest number of inputs. The (overall) Technical Efficiency (TE) is the product of Scale Efficiency (SE) and PTE [75,76] . TE is the efficiency measured under the CRS production frontier. While scale efficiency measures how far a unit strays from an optimal scale (which is the area where there are CRS in the relationship between outputs and inputs) [77,78]. As a result, it can be written as:

Technical Efficiency (TE), θ_CCR_ = Pure Technical Efficiency (PTE), θ_BCC_ x Scale Efficiency The Fig 2 illustrates geometrically the concepts of efficiency measurement in DEA Twenty nine percent (26 of 89, 29.21%) of the included studies measured the (overall) Technical Efficiency (TE) [77–102] and 26.97% (24 of 89) [75,76,103–124] used TE, PTE and SE in their evaluation. While the rest of the studies evaluated their efficiency by the combination of TE and PTE and some not stated clearly the type of efficiency measured (for details see Appendix C).

**Fig 2.**
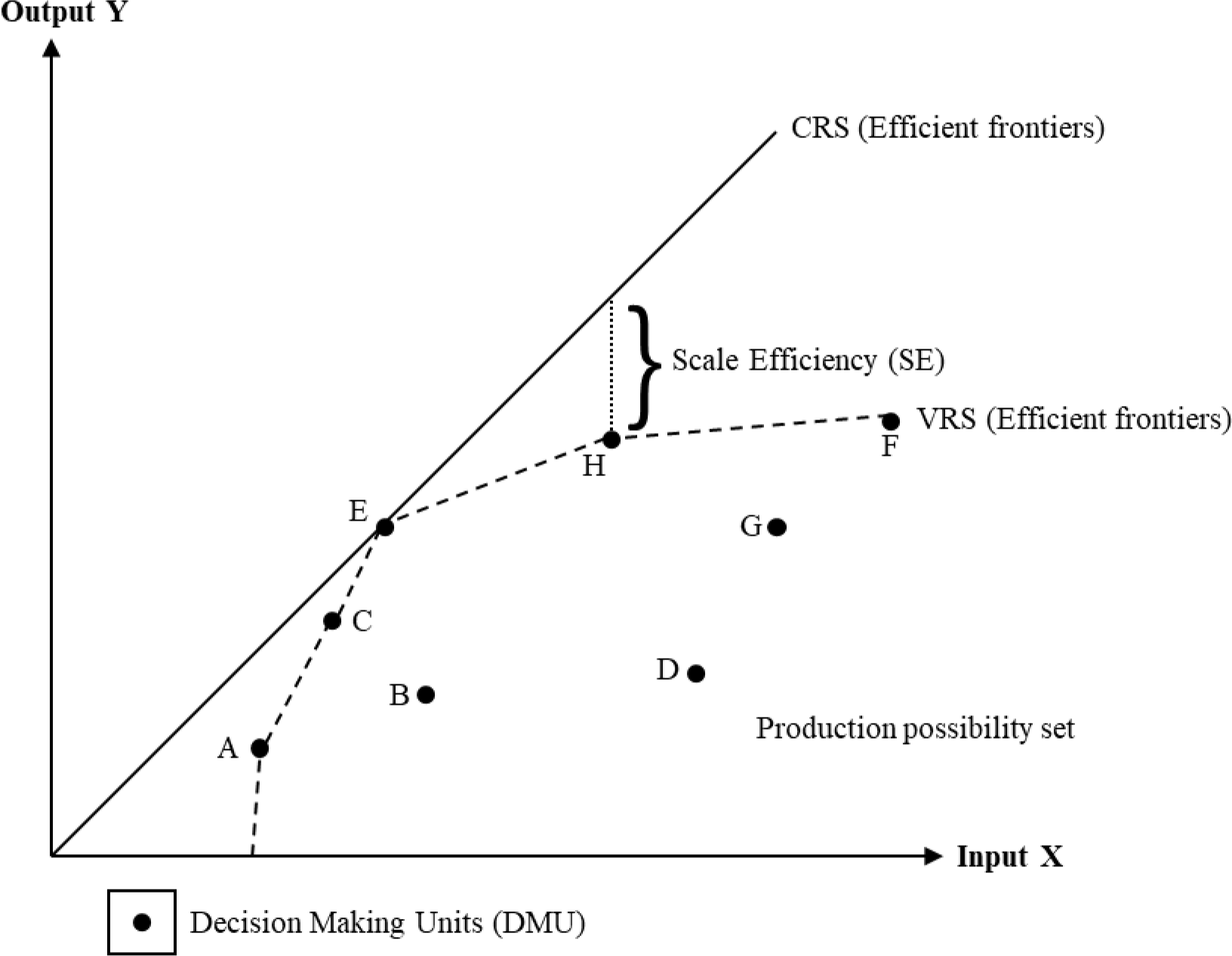
Concepts of efficiency measurement in Data Envelopment Analysis.

### 3.2 Model parameters

Four considerations must be specified by researcher or user in order to apply DEA: the model type, the technological assumption of the delivery process, the model orientation, and the input-output combination [113,122]. This model can be further analysed or extended via second stage analysis or integrated with other statistical analysis to attain better efficiency measurement, to explain the variation or difference in organization performance or to evaluate the productivity of organization over certain time [92,111,120]. The data type is also important, as it showed how the performance were analysed over certain time period.

#### 3.2.1 Model type

DEA has been applied to evaluating the performance of a wide range of entities engaged in a wide range of activities in a wide range of circumstances. Thus, lead to many models and extensions to explain the intricate and frequently unknowable relationships between the various inputs and outputs that are involved in organization activities or productions [107,125]. The models described as basic DEA model and extensions model. While others detailed the model as Radial, Non-Radial and Oriented, Non-Radial and Non-Oriented, and Radial and Non-Radial [23,126]. The majority of the research that were included used Radial DEA models (72 of 89, 80.90%) [46,48–52,54–62,65–77,79,80,82–86,89–94,96–100,102–116,118–124,127–130]. BCC, CCR or combination of both models were the specific model utilized in measuring efficiency via radial change of input and output values. In contrast, only seven studies (7.87%) [47,53,63,64,95,101,131] used Non-Radial and Oriented model. While four studies (4.49%) [81,87,88,125] used Non-Radial and Non-Oriented model. This Non-Radial model, instead of adhering to a proportionate change of input/output, deals with slacks directly. One study used Radial and Non-Radial [78] and one study applied combination of Radial with Non-Radial and Oriented model [117] to measure efficiency. While four studies [132–135] did not clearly state the model used in their paper (for details see Appendix D).

#### 3.2.2 Model orientation

Orientation indicates the input or output orientation in measuring efficiency. In other words, either output expansion or input decrease is the primary evaluation goal. More than half of the studies (55.06%, 49 of 89) [46–48,50–54,58,60,64–69,71–73,75–77,80,91–94,96,98,99,103–107,109–111,113–116,118,121,124,127,128,130,135] applied input-orientated DEA models. The researchers indicated that the choice of input orientation was made to be in line with the fact that the majority of healthcare units aim to minimize inputs given a goal level of output. In other words, the organization had little or no control over the output [46,51,110]. About a quarter of the studies (25.84%, 23 of 89) [49,55–57,59,61,62,70,74,79,82–86,102,108,112,117,119,122,123,134] expressed the opposite. The organization should me be able to produce more output given that the input is fixed and not easily increase. Thus, output-orientated DEA models are more appropriate in their settings [79,83,84]. Five studies (5.62%) [81,87,88,100,125] applied Non-Orientated DEA models, while three studies (3.37%) [95,101,129] used the combination of both input & output orientated DEA models. The rest of the studies [63,78,89,90,97,120,131–133] did not clearly state which orientation were used in their measurement (for details see Appendix E).

#### 3.2.3 Return to scale assumption

Regarding the return to scale assumption, about one third of the studies (35.96%, 32 of 89) [75,76,79,92,102–125,127–130] applied combination of both CRS and VRS assumptions in evaluating efficiency. The researchers seek to contrast the efficiency score measured in order to get better understanding of the organization. Also gave further insight on how they can use each assumption in improving their hospital services [102,109,114]. Another one third (32.58%, 29 of 89) [46–74] of the studies utilized VRS assumption and implied that the outputs of organizations (DMUs) changes significantly (increase/decrease) with inputs. While eighteen studies (20.22%) [77,80,82–86,89–91,93,94,96–101] relied on the CRS assumption that the outputs of their organizations (DMUs) vary (increase/decrease) in a manner similar to that of the inputs (for details see Appendix F).

#### 3.2.4 Input and output selection

It is essential for a meaningful evaluation to choose appropriate inputs and outputs. Finding characteristics that best describe the investigated process or production is one of the most crucial responsibilities. All relevant resources should be incorporated into the inputs, and the outputs should outline the organizations (DMUs) administrative goals [53,105]. The characteristics of suitable inputs and outputs can vary depending on the situation. Although it is crucial to select the appropriate inputs and outputs, data availability is also one of the considerations that need to be noted by the researcher. There are recommendations have been set up to assist researchers in locating suitable measures [77,132,134]. This will be discussed further as the main objective of this review.

There are classification of inputs and outputs to aid researchers in measuring efficiency. Namely, capacity-related, labour-related, and expenses-related. Or capital investment, labour and operating expenses. Some researcher further identified this classification into sub-categories. The outputs can be classified as inpatient services, outpatient services and effectiveness (quality) component. While other categorized outputs into activity-related (inpatient and outpatient) and quality-related (effectiveness dimension) [20,21,25,32]. In this review we classified and sub-classified the input & output as displayed in Table 1 and the details of each sub-classification frequency distribution and percentages as in Table 2 and Table 3.

**Table 1.** Input and Output Categories used in the studies.

| **Input** | **Percentage (%)** | **Output** | **Percentage (%)** |
| --- | --- | --- | --- |
| Capacity-related: | 30.06 | Production-related | 94.30 |
| Beds | 24.85 | Inpatients | 42.62 |
| Capital assets | 5.21 | Outpatients | 24.83 |
|  |  | Adjusted scores | 13.42 |
| Cost-related: | 15.34 | Combination | 12.08 |
| Total costs | 7.06 | Monetary | 4.70 |
| Medication & service costs | 5.21 | Imaging & Laboratory | 2.35 |
| Labour costs | 2.15 |  |  |
| Equipment costs | 0.92 | Quality-related | 5.70 |
|  |  | Patients | 72.22 |
| Staff-related: | 52.76 | Staffs | 27.78 |
| Doctors | 17.18 |  |  |
| Nurses | 13.19 |  |  |
| Clinical staffs | 10.74 |  |  |
| Non-clinical staffs | 7.98 |  |  |
| Combination | 3.68 |  |  |
| **Other specific output:** | 1.84 |  |  |

**Table 2.** Complete list of input used.

| **Input** | **Frequency** | **Percentage(%)** |
| --- | --- | --- |
| Capacity-related: |  |  |
| Number of general beds | 73 | 74.49 |
| Number of facility types (area, space) | 10 | 10.20 |
| Number of medical equipment | 5 | 5.10 |
| Number of acute beds | 4 | 4.08 |
| Adjusted beds value (ratio, log value) | 4 | 4.08 |
| Number of total assets | 2 | 2.04 |
| Cost-related: |  |  |
| Total operating costs | 14 | 28 |
| Fixed costs | 7 | 14 |
| Service costs | 5 | 10 |
| Consumable costs | 5 | 10 |
| Total labour costs | 4 | 8 |
| Combination | 4 | 8 |
| Medication costs | 3 | 6 |
| Clinical staff costs | 2 | 4 |
| Capital costs | 2 | 4 |
| Beds cost | 2 | 4 |
| Non-clinical staff costs | 1 | 2 |
| Medical equipment costs | 1 | 2 |
| Staff-related: |  |  |
| Number of Specialist, Physician, Doctor, GP, Dentist | 38 | 22.09 |
| Number of Nurses, Midwives, Nursing staffs | 30 | 17.44 |
| Number of Medical staffs | 23 | 13.37 |
| Number of Non-clinical staffs | 18 | 10.47 |
| Full Time Equivalent Specialist, Physician, Doctor, GP, Dentist | 16 | 9.30 |
| Full Time Equivalent Nurses, Midwives, Nursing staffs | 11 | 6.40 |
| Number of combination of staffs | 9 | 5.23 |
| Full Time Equivalent Non-clinical staffs | 8 | 4.65 |
| Full Time Equivalent Medical staffs | 6 | 3.49 |
| Number of Allied Health staffs | 5 | 2.91 |
| Ratio of Doctors | 2 | 1.16 |
| Ratio of Nurses | 2 | 1.16 |
| Full Time Equivalent combination of staffs | 2 | 1.16 |
| Number of combinations of Medical staffs | 1 | 0.58 |
| Combination of staffs Log value | 1 | 0.58 |
| Others: |  |  |
| Number of admissions | 1 | 16.67 |
| Average Length of Stay | 1 | 16.67 |
| Annual revenue | 1 | 16.67 |
| Discharge Log value | 1 | 16.67 |
| Inpatient discharge rate | 1 | 16.67 |
| Population | 1 | 16.67 |

**Table 3.** Complete list of output used.

| Output | Frequency | Percentage(%) |
| --- | --- | --- |
| Production-related: |  |  |
| Number of Outpatients | 50 | 16.78 |
| Number of Inpatients (admission, discharge) | 33 | 11.07 |
| Total number of operations | 28 | 9.40 |
| Number of Inpatients | 23 | 7.72 |
| Number of Emergency Outpatients | 22 | 7.38 |
| Number of Inpatient days | 14 | 4.70 |
| Bed Occupancy Rate (BOR) | 12 | 4.03 |
| Inpatient adjusted value (Casemix, price, ratio) | 12 | 4.03 |
| Discharge adjusted value (Casemix) | 11 | 3.69 |
| Number of General & Emergency Outpatients | 10 | 3.36 |
| Average Length of Stay (ALOS) | 9 | 3.02 |
| Total revenue | 8 | 2.68 |
| Ratio adjusted value | 7 | 2.35 |
| Number of Daycare patients | 6 | 2.01 |
| Number of birth deliveries (normal, caesarean) | 6 | 2.01 |
| Number of laboratory & radiology (services, examinations) | 5 | 1.68 |
| Number of operations | 4 | 1.34 |
| Outpatient adjusted value (Casemix, price, ratio) | 4 | 1.34 |
| Number of Total patients | 3 | 1.01 |
| Number of Family Medicine outpatients | 2 | 0.67 |
| Number of Obstetric outpatients (ANC, PNC) | 2 | 0.67 |
| Number of emergency outpatient surgeries | 2 | 0.67 |
| Bed Turnover Rate (BTR) | 2 | 0.67 |
| Bed Turn Over Interval (TOI) | 2 | 0.67 |
| Number of surgeries (Minor, Major) | 2 | 0.67 |
| Log adjusted value | 2 | 0.67 |
| Score adjusted value | 2 | 0.67 |
| Operating income | 2 | 0.67 |
| Inpatient income | 2 | 0.67 |
| Number of diagnostic (visits, procedures) | 2 | 0.67 |
| Number of Medical Outpatients | 1 | 0.34 |
| Number of Allied Health Outpatients | 1 | 0.34 |
| Average number of admission & discharge | 1 | 0.34 |
| Number of Special patients | 1 | 0.34 |
| Number of total examinations | 1 | 0.34 |
| Discharge adjusted value (Casemix) | 1 | 0.34 |
| Death adjusted value (Casemix) | 1 | 0.34 |
| Outpatient revenue | 1 | 0.34 |
| Examination revenue | 1 | 0.34 |
| Quality-related: |  |  |
| Mortality rate (infant, adult, specific diseases) | 9 | 50.00 |
| Revisit rate (outpatient, emergency) | 3 | 16.67 |
| Number of students | 2 | 11.11 |
| Patient's satisfaction score | 1 | 5.56 |
| Staff's satisfaction score | 1 | 5.56 |
| Number of medical inquiries | 1 | 5.56 |
| Management's score | 1 | 5.56 |

##### 3.2.4.1 Capacity-related input

Most frequently, the size, capacity, or functioning of a hospital as a health service is determined by the number of fully staffed hospital or operational beds. 75 of the 89 (84.27%) studies included the number bed (i.e., general, ICU, special) as input in their analysis [48–55,57–62,64–74,76,78–93,95–101,103–109,112–116,118,119,121–125,128–131,133–135]. Seven of these 75 studies (9.33%) utilized variation of multiple beds as their inputs either by type of bed, bed cost, or ratio of bed [61,66,71,78,98,101,129]. While 12 of these 75 (16.00%) studies combine both beds and capital assets as their capacity-related input [70,78,85,89,90,101,103,106,109,122,125,133]. One study only used capital assets as their input but combine with cost-related assets, this may explain why this study did not use beds as part of input like other studies [56]. The most prevalent capacity-related input in the studies was the number of general beds, followed by the number of facility types and finally the number of medical equipment.

##### 3.2.4.2 Cost-related input

Among the listed research, cost-related inputs were the least utilised inputs. Out of the 89 studies, only 31 (34.83%) applied in the studies. Three of these 31 studies, specifically used cost-related inputs only in their research [63,102,120]. While majority of these 31 studies (90.32%) utilized combination of either capacity-related inputs and staff-related inputs in their research [46–48,51,56,62–64,70,75–79,83,91,94,95,101,102,109,111,112,114,116,120,125,129,130,132,133]. The most often used cost-related input in the research was the total operating cost, followed by the fixed costs, finally both service costs and consumable costs.

##### 3.2.4.3 Staff-related input

Almost all of the studies utilized staff-related inputs (93.26%, 83 of 89) [46–55,57–62,64–90,92–119,121–125,127–131,134,135]. Out of these 83 studies, two studies combined used the number of staff (staff-related) and the labour cost (cost-related) their input [75,78]. The six studies that did not included staff-related input, substitute cost-related (either labour cost or operating cost) as their proxy in their analysis [56,63,91,120,132,133]. The staff-related input values varied among the observed papers. Most of the studies used arithmetic number (actual), followed by Full Time Equivalent and ratio of specific value. The number of doctors, followed by number of nurses and number of clinical staffs, was the most prevalent staff-related input in the studies.

##### 3.2.4.4 Production-related output

Looking at the output, almost all of the studies used production-related output (98.88%, 88 of 89) [46–92,94–125,127–135]. Eight studies applied combination of production-related output with quality-related output [61,73,84,87,95,98,128,133]. The number of outpatients were the most prevalent production-related output among the studies. The number of inpatients (admission, discharge) were the second most prevalent, followed by the total number of operations and the number of inpatients.

##### 3.2.4.5 Quality-related output

The application of quality-related outputs was varied and were not prominent as the production-related output. Nine studies used quality-related output [61,73,84,87,93,95,98,128,133]. One study specifically applied only quality-related in their research, which match with the researcher objective [93]. The most often applied quality-related output was mortality rate (infant, adult, specific diseases), followed by revisits rate (outpatients, emergency) and number of students.

#### 3.2.5 Extended analysis & data type

Among the studies, 80 (89.89%) conducted extended analysis in their research [46–56,58–64,66–69,71–77,79–94,96–98,101–116,118–125,128–135]. The type of data applied were almost equally distributed among the studies 51 (57.30%) [47,49,52,58–60,67,70–73,75–77,79,80,83–85,88,91,92,94,96,97,100,101,103,105–109,111,112,114–116,120–122,124,125,127,129–135] used panel data and 38 (42.70%) [46,48,50,51,53–57,61–66,68,69,74,78,81,82,86,87,89,90,93,95,98,99,102,104,110,113,117–119,123,128] used cross sectional data in their research. Forty extended analyses were identified within the included studies. Each study may apply one or multiple extended analysis (some mentioned as “stages”). Out of 80 articles that applied extended analysis, 46 studies integrated two or more extended analysis in their DEA measurement [46,47,51–54,58,60,63,64,67,68,71,73–75,77,79,80,83–86,88,91–94,96,101,102,107–109,114–116,119–121,128,130,131,133–135]. The maximum number of extended analyses observed within the studies were five [67,92,94]. The main extended analysis used in hospital efficiency assessment was regression analysis (29.11%, 46 of 158) followed by production function analysis (16.46%, 26 of 158), statistical analysis (15.82%, 25 of 158) and resampling methods (15.82%, 25 of 15). The complete list of the specific analysis to each classification is presented in Table 4.(for details see Appendix B)

**Table 4.** Complete list of extended analysis.

| Extended analysis | N | % | Extended analysis | N | % |
| --- | --- | --- | --- | --- | --- |
| Regression analysis | 46 | 29.11 | Production function analysis | 26 | 16.46 |
| Tobit regression | 26 | 16.46 | Malmquist Productivity Index | 21 | 13.29 |
| Ordinary Least Squares regression | 11 | 6.96 | Window analysis | 3 | 1.90 |
| Truncated regression | 5 | 3.16 | GML<sup>a</sup> Index | 1 | 0.63 |
| Generalized Estimating Equations regression | 1 | 0.63 | gMMP<sup>b</sup> Index | 1 | 0.63 |
| Linear regression | 1 | 0.63 |  |  |  |
| Multinomial Logit regression | 1 | 0.63 | Resampling methods | 25 | 15.82 |
| Beta regression | 1 | 0.63 | Bootstrap method | 24 | 15.19 |
|  |  |  | Monte Carlo simulation | 1 |  |
| Statistical analysis | 25 | 15.82 |  |  |  |
| Mann–Whitney U test | 5 | 3.16 | Performance measurements | 13 | 8.23 |
| Wilcoxon signed-rank test | 4 | 2.53 | Benchmarking method | 7 | 4.43 |
| Kruskal-Wallis test | 4 | 2.53 | Stochastic frontier analysis | 2 | 1.27 |
| F-test | 2 | 1.27 | Isodata analysis | 1 | 0.63 |
| T-test | 2 | 1.27 | Super efficiency analysis | 1 | 0.63 |
| Paired T-test | 1 | 0.63 | Fitting adjustment | 1 | 0.63 |
| Central Limit Theorem | 1 | 0.63 | Pabon Lasso technique | 1 | 0.63 |
| Li-test | 1 | 0.63 |  |  |  |
| Chi-square test | 1 | 0.63 | Corelation analysis | 11 | 6.96 |
| Repeated measures ANOVA | 1 | 0.63 | Spearman's rank correlation | 8 | 5.06 |
| Theil index | 1 | 0.63 | Pearson's correlation | 2 | 1.27 |
| Unpaired T-test | 1 | 0.63 | Grey's correlation | 1 | 0.63 |
| Univariate analysis | 1 | 0.63 |  |  |  |
|  |  |  | Clustering analysis | 8 | 5.06 |
| Matching methods | 3 | 1.90 | Cluster analysis | 6 | 3.80 |
| Propensity score matching | 3 | 1.90 | k-means clustering | 1 | 0.63 |
|  |  |  | MST-kNN clustering algorithm | 1 | 0.63 |

<sup>a</sup> global Malmquist-Luenberger
<sup>b</sup> generalized metafrontier Malmquist productivity

### 3.3 Approach in selection of input and output

In selecting input and output for DEA with regards to hospital, various approaches or method adopted by the researchers. All the 89 studies used previous study or literature review as a sole basis or part of their method in selecting input and output. Few studies precisely mentioned using local DEA efficiency study at their respective country as the reference for input and output selection. Some of the studies used combination of approaches. 63 studies (70.79%) used only literature review as their approach in selecting the input and output [46–48,50–55,59,60,62,64–78,81,82,85,86,89–92,94,96–98,101,103,105,108,110,111,113,115–122,124,125,127,128,130,132–135]. While the rest used literature review in combination with other approaches. The combination was varied among the studies.

The dominant combination approach identified was literature review & data availability (13.48%, 12 of 89) [56,57,79,83,104,106,107,112,114,123,129,131], followed by literature review & systematic method (5.62%, 5 of 89) [58,61,84,102,109] and literature review & DMU limitation (5.62%, 5 of 89) [49,80,87,95,99]. The maximum combination was four, which one study mentioned in their article [88].The complete list of the specific approach presented in Table 5

Classification of approach in selection of input and output.

**Table 5. Classification of approach in selection of input and output**
| Approaches | N | % |
| --- | --- | --- |
| Literature review | 63 | 70.79 |
| Literature review | 60 | 67.42 |
| Local studies literature review | 3 | 3.37 |
| Literature review & data availability | 12 | 13.48 |
| Literature review & data availability | 11 | 12.36 |
| Local studies literature review & data availability | 1 | 1.12 |
| Literature review & systematic methods | 5 | 5.62 |
| Literature review & Delphi method | 2 | 2.25 |
| Literature review & Promethee method | 1 | 1.12 |
| Literature review & Bibliometric analysis | 1 | 1.12 |
| Literature review & Variance filter analysis | 1 | 1.12 |
| Literature review & DMU limitation | 5 | 5.62 |
| Literature review & expert judgement | 3 | 3.37 |
| Literature review, data availability, expert judgement & DMU limitation | 1 | 1.12 |

## Discussions

Anyone unfamiliar with the subject will find the DEA universe’s size to be intimidating. It is practically impossible to read every previous study to gain knowledge from their experiences, even when limiting the literature to healthcare applications. To accomplish the goals of this study, 89 studies underwent thorough scrutiny. Nunamaker published the first health application utilising DEA to investigate nursing services, and Sherman published the second DEA paper to assess the medical and surgical departments of seven hospitals [136,137]. Since then, Over the past forty years, DEA applications in health have evolved. The calibre of the studies has increased as access to resources and information technology has advanced [23,25,138,139].

### 4.1 Approaches taken by researchers in selection of input and output of DEA for hospital efficiency measurement

DEA is frequently used to assess the relative effectiveness of a group of institutions like businesses, hospitals, universities, and government agencies. In conventional analysis, such judgements might take many different forms. In hospitals setting, production of healthcare is distinct from manufacturing. Raw materials are physically transformed into finished goods in a conventional factory. There is no participation or co-production because the customer is not present. It is challenging to identify the appropriate variables because patients are involved in the process. Furthermore, in healthcare it is not just about performance and efficiency, the effectiveness (quality component) is equally important [87,93,128]. Although there is no standard set of input and output in DEA studies, there are several guidelines, analytic procedures or principles to help researchers choose the best variables in their study [140–144].

#### 4.1.1 Literature review

A literature review is still used today and is thought to be one of the most effective techniques to place a study within the body of knowledge. Be it in the form of narrative review, rapid review, scoping review or systematic review. It acts as a foundation or building block for knowledge advancement, theory development, and identify areas of improvement [145–147]. Literature reviews were the most prevalent approach for selection of input and output for hospital efficiency study using DEA. Our reviews showed that all studies applied literature review as sole approach or part of combination approach. None of the studies stated exactly on the methodology of literature review done except few studies that define they only taken studies from their local country [50,86,105,123]. The reason for researchers to specify this is one of their objectives was to compare their findings with local previous studies. DEA is a non-parametric technique that relies entirely on the observed input-output combinations of the sampled units and do not necessitate any presumptions regarding the functional structure of the link between inputs and outputs [77,115]. Thus, this gives the advantage for researchers to select input and output based on literature review as DEA will measure the efficiency value which depend on the study objective even though it might give a less significant value [140,143].

#### 4.1.2 Data availability

DEA relies on homogeneity of the unit’s assessment. The DMU are assumed to be producing similar activities or product with similar resources and technology under the same environment. Thus, a common set of similar input and output can be defined . With large datasets in the hospitals, there are needs some consideration to be done. This includes, the data quality, data availability, data scale and types of data [140,142]. Within the studies it is found that, although the researchers applied literature review in selection of input and output, they mentioned it is subjected to data availability. Few methods identified on how the researchers overcome this limitation. First, specifically mentioned that the study only collected data which is available within their scope [88,104,131]. Second, some study omitted the DMU with incomplete data and focus their analysis on DMU with complete data [79,106]. This is one of the advantages of DEA, as DEA only measure the relative efficiency or the production frontier of the units include in the analysis. Third, the researcher applied DEA within specific period of data availability to ensure the desired input and output are complete [56,114]. There is method in dealing with missing of incomplete data in DEA, but non-of the studies reviewed applied of mentioned on it [148,149].

#### 4.1.3 Systematic method

In determining the input and output, researchers can list as many as possible factors. This may result in two major issues. One there will be a long list of input and output and second, if there is limited number of DMU the discriminating power for DEA to measure the efficiency will be affected [140,143]. It is important to choose the important variables and at the same time able to measure the efficiency accordingly. Researchers has been incorporating systematic procedure to judge this process. In the review, four systematic approaches have been identified which are Delphi method, Preference Ranking Organization METHod for Enrichment of Evaluations (PROMETHEE), Bibliometric analysis, and Variance filter [49,58,61,102,109]. The Delphi method is a methodology used in research models that is centred on gathering the most trustworthy consensus of expert opinion for challenging situations. This forecasting method was first presented in the 1950s by Olaf Helmer and Norman Dalkey of the Rand Corporation. It is based on the responses to multiple rounds of questionnaires sent to a panel of experts [150,151]. The Delphi method is a well-known method for researchers to use in measuring efficiency using DEA in various area [152–154]. In 1982, the PROMETHEE method was created for the first time, and in 1985 it underwent further development [155–157]. It is currently regarded as a widely used and usable Multiple Criteria Decision Aid tool including application with DEA [158–161]. In comparison to many other MCDM methods, PROMETHEE is a ranking system that is thought of as being simple in conception and computation. It includes weights indicating the relative importance of each criterion as well as a preference function linked to each criterion. One of PROMETHEE’s key applications, the ability to select, is used to help the decision-maker select the best possibilities for assessing hospital performance. This gives the ability for researchers to integrate PROMETHEE in DEA application [162–165]. The term bibliometric was coined by Pritchard in 1969. It is an “application of mathematics and statistical methods to books and other media of communication”. In other words, bibliometric analysis evaluates the properties of bibliographic information or metadata from database or collection of documents. With a purpose to increase knowledge of the topic being studied [166,167]. Researchers applies bibliometric analysis for various objectives, to identify new trends in journal performance, collaborative styles, and research components; to lay the groundwork for the new and significant advancement of a field; and to making sense of massive amounts of unstructured data in a systematic manner in order to interpret and map the cumulative scientific knowledge and evolutionary nuances of established domains [168,169]. Thus, the usage of bibliometric analysis is applicable in identifying the input and output in DEA studies with regards to the field of healthcare especially hospital. Variance filter is one of the filter methods in Feature Selection. It is the process of deciding which traits are most crucial and keeping them. It helps to lessen noise, lower the model’s computational expense, and occasionally boost model performance. In this case which in the crucial input and output listed by the researchers [170,171]. By using the Variance filter (Feature Selection), researchers have the ability to remove variables (input or output) which have little or no impact in the measurement of DMUs efficiency. This method is well accepted in the application of DEA studies [102,172–174].

#### 4.1.4 Expert judgement

The refinement of variables selection (input and output) can be decided by the judgement of expert which can be the researcher, the stakeholders or the decision makers. The value judgment can be said as “logical constructs used in efficiency assessment research that reflect the Decision Makers’ (DM) preferences during the efficiency assessment procedure”. This includes the decision to omit variables or the put zero weight in the variable [143,175]. This is again the ability of DEA to measure efficiency based on the needs and requirement of the decision makers. Although it can lead to problem such as bias in selection, omission of input or output that may have significant impact on the efficiency measurement, or putting the wrong weight for input and output. The incorporation of expert or value judgment are motivated by the objective of researchers. There are advantages and disadvantages with respect to the application of DEA. It is important to understand the managerial and statistical implication of applying value judgement in the selection of input and output [176].

### 4.2 Common DEA model parameters in hospital efficiency evaluation

#### 4.2.1 Model type

The application of DEA has been undergoing extensive developments and evolution over the time. Thus, numerous models exist to measure efficiency from the basic model to the very specific application of DEA. In this review we found that 80.90% of the included studies applied Radial DEA models. Which includes BCC model (Banker, Charnes and Cooper), CCR model (Charnes, Cooper and Rhodes), and combination of both models. These findings are similar with some of the previous review in healthcare setting [17,21,33]. A Radial DEA model means the major concern is a proportionate change in the input/output values; as a result, slacks (input excesses and output shortfalls still present in the model) are ignored or treated as optional. Although there are limitations with radial model, researchers still use this model as it is a basic model, simple to understand and easier to apply without much requirement on the production criteria of the DMUs [48,80,177].

#### 4.2.2 Model orientation

The selection of DEA orientation depends on several factors or arguments. It can be what the decision maker can control more effectively, what is the nature of the production, or what is the researcher objective from the model [25,32,178]. Healthcare organizations or hospitals in general have limited or less control over their outputs. But it does not mean a DEA efficiency evaluation in hospital must be in input orientation. Our review, found that 55.06% of the articles used input-oriented DEA models. Previous studies also shown similar findings albeit in different proportion [17,20,21]. This can be seen as researchers or hospital managers consider that minimising inputs given a goal level of outputs is more suited for measuring hospital efficiency since hospital have limited control over their outputs.

#### 4.2.3 Return to scale assumption

Since the development of the two fundamental models, there has been debate on which is better (and, consequently, whether to assume CRS or VRS). Hospital managers seek the best method evaluations to measure which input or output are improving their organization’s efficiency. The selection of return to scale with regards to hospital depends on the size of hospital [65], the organization factors [76,91], the flow of process of input and output [111,127], technological involvement [82,122], or other related factors. It is crucial to keep in mind that the adoption of an improper return to scale may result in an excessively constrained region of the search for effective DMUs. Thus, the researcher can consider to explore both assumptions to understand the implications of using either one [179]. In the review, we observed most of the studies applied both assumption for comparison reason (35.96%) and followed by VRS assumption (32.58%). In past review, there is a shift towards using CRS to VRS assumption in application of DEA [20,21,33]. It may be said that the majority of assessments of hospital efficiency were made with of view economies of scale exists and that they considered the production function of healthcare to have non-proportionality shifts between inputs and outputs.

#### 4.2.4 Inputs and outputs choice

We already covered the main objective of this review to look at the approaches taken by researcher in selection of their inputs and outputs. The selection of appropriate inputs and outputs is essential for the analysis of DEA to be useful, and we cannot emphasise this point enough. As many previous reviews stated, the quality and quantity of these indicators play a major role in the DEA efficiency analysis [15–18,20,21,30,33]. We observed in term of inputs, majority of the studies (52.76%) used staff-related input as part of the variables in efficiency measurement. This is not surprising considering human resource plays an important role in any organization including hospital. This finding is similar to previous study of DEA and even in other performance related studies on healthcare service [180,181]. The unit of analysis of staff-related factors depends on how the organization functioning. In the review mainly we noticed to types either actual number of staff or full time equivalent. The spectrum of staff types was various from the clinical staff to non-clinical staff (Appendix A and B). If we looked at the sub-type of inputs, the number of general beds had the highest prevalence (74.49%). Hospital beds is the basic capital-related input for a hospital. It is one of the key indicators that use for hospital performance, hospital capacity, hospital capability even in comparing health service between country [182–184]. As for the outputs, in general we observed most researchers applied production-related outputs compare to quality related. This occurrence may be said due to it is easier to measure production-based data and give the stakeholders a relatively target to improve upon it. While effectiveness (quality) is not something that healthcare managers would trade-off to improve the efficiency [25,185]. In the review, the number of inpatients, number of our patients and number of operations were the common outputs applied. The widespread use of these variables was no surprised, as it is the bread and butter of hospital services.

## Limitation & conclusion

This is the first in-depth systematic review examining the approach in input-output selection of DEA efficiency measurement in hospital. As far as we know, no research has previously examined this. That is the main purpose of this systematic review to provide an overview of the existing approaches. In addition to serve as an update on DEA models that are currently applied for hospital efficiency evaluation. Eighty-nine articles were reviewed and assessed thoroughly with the mentioned objectives. Literature review was dominantly utilized as an approach for selection of inputs and outputs in DEA. Researchers used literature review as a sole method or combined with other approaches to make the selection more robust and vigorous. As the selection of variables in DEA study are very important and may cause different results in efficiency measurement [140]. There is no specific approach or methods that can be point out in selection of variables (input-output) in DEA. That is the advantage and also weakness of DEA [186–188]. It gives the ability for researchers and stakeholders to evaluate efficiency of their organization based on their preference but they need to be aware of the limitation of DEA and potential pitfall [140,143]. This review focus on approaches in hospital settings, this may limit our findings. There might be other approach or method that is use to choose input and output for DEA study in another field or approaches based on other perspectives [189]. The approach in input-output selection should be identified given the ongoing aim of researchers and healthcare professionals to improve healthcare efficiency assessment. It is crucial to consider past, present, and potential developments on this subject in the DEA literature because it plays a vital role in DEA studies. As for the DEA models parameters, there is no evidence of one best model or one fits model as almost all models had been utilized more than twice (Appendix A and B). Our review provides some guideline and methodological principle on DEA study based on established literature. This hopefully will shed some light to hospital managers, healthcare workers, policy makers even students on evaluation of efficiency by DEA.

## Data Availability

All relevant data are within the manuscript and its Supporting Information files.

## Registration and protocol

This study was registered at OSF Registries (https://osf.io/registries) all information regarding the registration and study protocol can be access at https://osf.io/nby9m or https://osf.io/e7mj9/?view_only=53deec8e6c6946eeaf0ea6fe2f0f212a.

## Author Contributions

MZZ, AAN and AMMR: conception and design of study. MZZ: acquisition of data. MZZ, AAH and MIA: analysis of collected data. MZZ, AAN, AMMR: interpretation of data. MZZ, ANN, AMMR, AAH, MIA, ZZ and AFAB drafting of paper and/or critical revision. MZZ had primary responsibility for final content. All authors read and approved the final manuscript.

## Data Availability Statement

The original contributions presented in the study are included in the article/supplementary material, further inquiries can be directed to the corresponding author/s.

